# Sex-Specific Associations of Carotid Plaque Calcification Subtypes with Stroke, Transient Ischemic Attack, and Retinal Infarction

**DOI:** 10.64898/2026.09.14.26363078

**Authors:** Henna Komi, Petra Ijäs, Krista Nuotio, Laura Mäkitie, Mikko I. Mäyränpää, Mohammadreza Shoghli, Riikka Tulamo, Pirkka Vikatmaa, Perttu J. Lindsberg, Lauri Soinne, Suvi Koskinen, Juha Sinisalo, A. Inkeri Lokki

## Abstract

**Background:** Calcification in carotid plaques has been suggested to represent a stabilizing plaque feature, but previous findings are conflicting. Whether histologically defined calcification subtypes are associated with distinct cerebrovascular event phenotypes and whether these associations differ by sex remain unclear.

**Methods:** In this prospective Helsinki Carotid Endarterectomy Study 2, carotid plaques from 483 patients (328 men) undergoing carotid endarterectomy were analyzed histologically using AI-based quantification of sheet, nodular, and total calcification (%). Calcification measures were compared with patient characteristics, plaque morphology, CTA-based calcium score, and neurological symptom subtypes (asymptomatic (n=167), stroke (n=132), transient ischemic attack (TIA) (n=79), retinal infarction (n=24), and amaurosis fugax (n=81)), with sex-stratified analyses.

**Results:** Women had more plaque calcification than men (median total calcification, 15.8% versus 10.8%; p=0.002). In the overall cohort, symptomatic plaques showed lower total and sheet calcification than asymptomatic plaques, an association driven by men. Among male patients, stroke-associated plaques had less sheet calcification than asymptomatic plaques (median, 3.4% versus 8.4%; p<0.001). In women, plaques associated with TIA showed higher total calcification than stroke-associated plaques (median, 22.7% versus 13.5%; p=0.024). Retinal infarction was associated with higher nodular calcification than stroke in the overall cohort (median, 9.6% versus 1.2%; p=0.004). Histological total calcification correlated with CTA-based calcium score (p<0.001), and radiological calcification showed corresponding associations with neurological symptom type.

**Conclusions:** Overall, symptomatic plaques showed lower total and sheet calcification than asymptomatic plaques. In men, low total and sheet calcification were associated with stroke, whereas higher nodular calcification was associated with retinal infarction. In women, high total calcification was associated with TIA. These findings suggest that although calcification appears to be a stabilization plaque feature, its clinical implications depend on its morphological subtype and vary according to sex and neurological symptom subtype.

## Introduction

Atherosclerosis is a multifactorial disease characterized by the accumulation of lipid-laden plaques within the arterial wall and remains the leading cause of morbidity and mortality worldwide.^1^ Carotid atherosclerosis affects more than 1.3 billion individuals globally and is a major contributor to various cerebrovascular events. Carotid atherosclerosis accounts for an estimated 10–20 % of ischemic strokes and transient ischemic attacks (TIAs), and for an even greater proportion of retinal infarctions and amaurosis fugax.^2–4^

Carotid endarterectomy (CEA) reduces the risk of subsequent cerebral events in patients with carotid atherosclerosis selected based on symptom status and degree of stenosis.^5^ In addition to its clinical benefits, excised plaque material provides a unique opportunity for histological analyses, enabling deeper insights into the pathophysiology of atherosclerosis.^6^

The role of calcification in carotid plaques has been investigated in various clinical setups, yielding conflicting results. Most studies associate plaque calcification with increased plaque stability,^7,8^ whereas radiologically defined spotty calcification patterns have been linked to cerebral ischemic events.^9^ Histologically, plaque calcification can be classified into sheet calcification and nodular calcification. Sheet calcification develops through the coalescence of smaller calcium deposits into larger, homogenous calcium plates. In contrast, nodular calcification is characterized by irregular calcified nodules surrounded by fibrin deposits and has been proposed to have a thrombotic origin, as nodular calcification has been observed within old thrombi.^6,10^

Sex differences in atherosclerotic disease have gained increasing attention in recent years. Disease progression differs between the sexes, with men developing atherosclerotic disease 10–20 years earlier than women.^11,12^ This difference has been largely attributed to the protective effects of estrogen, as atherosclerotic progression accelerates after menopause.^13,14^ Consequently, middle-aged men have a higher risk of stroke, whereas in older age groups, postmenopausal women exhibit a higher risk than age-matched male counterparts.^15,16^ At plaque level, men have been shown to have larger carotid plaques and more frequently exhibit vulnerable plaque features—such as intra-plaque hemorrhage, lipid-rich necrotic core, and thin fibrous cap— compared with women. However, whether female plaques exhibit a higher relative degree of calcification remains unclear.^17–20^

Previously, one histological study compared carotid plaques from patients with cerebral ischemic events and retinal ischemia, demonstrating that plaques associated with retinal infarction contained fewer vulnerable features but more calcification than plaques associated with cerebral events.^21^ Another study reported that plaques from stroke patients exhibited more unstable features than those from TIA patients.^22^ Neither study included sex-stratified analyses.

To our knowledge, no previous study has comprehensively evaluated histologically defined carotid plaque calcification subtypes in relation to specific neurological event phenotypes with sex-stratified analyses. We investigated the associations of sheet and nodular calcification with baseline characteristics, plaque morphology, and distinct neurological symptom types in patients undergoing carotid endarterectomy. To support potential clinical translation, histological calcification measures were also compared with CTA-based radiological calcification assessment.

## Methods

### HeCES2 study description

The Helsinki Carotid Endarterectomy Study 2 (HeCES2) is a single-center, prospective cohort study comprising 500 consecutive patients undergoing carotid endarterectomy (CEA).^23^ Following referral, the decision to perform CEA was based on the European Stroke Organisation guidelines of the time.^24^ Patient recruitment took place between October 2012 and September 2015 at Helsinki University Central Hospital, Finland.

The cohort included both symptomatic (n=316) and asymptomatic (n=167) carotid stenosis patients and the relationship between the plaque and symptom was also determined in more detail. Plaque symptom status was independently assessed by two neurologists (K.N., P.I.), who determined whether the patient had experienced ischemic cerebrovascular symptoms attributable to the carotid plaque, or whether the plaque was asymptomatic. In cases of uncertainty regarding symptom etiology, a multidisciplinary panel of stroke neurologists (P.I., K.N., L.S., and P.J.L.) reached a consensus decision.

### Radiological calcification quantification (CTA-based calcium score)

Of the whole study cohort, 477 patients underwent pre-operative carotid CTA, 22 magnetic resonance angiographies, and one patient with contraindications for both underwent duplex sonography only. For diagnostic consistency, we included only CTA examinations in the radiological calcification quantification. The degree of carotid artery stenosis (%) was determined according to the North American Symptomatic Carotid Endarterectomy Trial (NASCET) criteria.^25^

Based on CTA, a semi-quantitative radiological calcium score was assigned to all plaques, as previously described.^26^ Briefly, radiological assessment was performed using the Impax workstation (AGFA Impax version 6.6.1.5003). The calcification burden of the internal carotid artery (ICA) was evaluated along the typical operative extent of CEA. The radiological ICA calcification score was defined as follows: (0) no calcification within the plaque (allowing 1–2 spot-like calcifications <0.5 mm in diameter); (1) numerous small calcifications within the plaque area; (2) fully calcified plaque with uniform bulky calcification as the dominant feature.

### Plaque assessment and AI-driven calcification quantification

Of the 500 consecutive patients, plaque specimens from 483 patients were successfully obtained and included in the analyses. After tissue processing including decalcification of the plaque, ^23^ hematoxylin and eosin (HE) stained plaque sections were digitized and plaque calcification was quantified using a cloud-based deep learning image analysis platform (Aiforia Create, Aiforia Technologies Oy, Helsinki, Finland, https://www.aiforia.com/) (Supplementary Figure S1). The development, training, and validation of the deep learning algorithms used to quantify nodular and sheet calcification in atherosclerotic plaques have been described previously.^27^ The algorithm segmented sheet and nodular calcification on digitized HE-stained plaque sections, and calcification areas were expressed both as absolute area (mm²) and as proportions of total plaque area.

### Statistical analyses

Normally distributed continuous variables are presented as mean ± standard deviation (SD) and were compared using Student’s *t*-test. Non-normally distributed variables are presented as median with interquartile range (IQR) and were compared using the Mann–Whitney U test. Associations between categorical variables were assessed using the chi-square test.

Differences in plaque calcification between asymptomatic and symptomatic patients were assessed using logistic regression models with each calcification measure entered separately as an explanatory variable. Odds ratios were reported per 10-% increase with 95% confidence intervals (CIs). Models were adjusted for sex, age, diabetes, coronary artery disease, smoking, lipid-lowering treatment, antithrombotic therapy, renal dysfunction, stenosis severity and prior stroke or TIA. Calcification-by-sex interaction terms were fitted to assess whether associations differed between male and female patients.

Differences across neurological symptom subtypes were evaluated using the non-parametric Kruskal–Wallis test, followed by Dunn post hoc test with Bonferroni correction. Neurological symptom types were categorized as follows: 0 = asymptomatic, 1 = stroke, 2 = retinal infarction, 3 = hemispheric transient ischemic attack (TIA), and 4 = amaurosis fugax.

The correlation between histological calcification measures and the radiological CTA-based semi-quantitative ICA calcification score was evaluated using Spearman’s correlation analysis.

Associations between symptom status and CTA-based calcium score (0–2) were assessed using ordinal logistic regression, with asymptomatic patients serving as the reference group. Odds ratios (ORs) and 95% confidence intervals (CIs) were estimated in the overall cohort and separately in men and women. The proportional odds assumption was verified using the Brant test. A two-sided p-value <0.05 was considered statistically significant.

In all analyses, statistical significance was defined as a two-sided *p* value <0.05. All statistical analyses were performed using R (RStudio version 2023.12.1) and SPSS version 29.0.0.0.

## Results

### Study participants

Baseline characteristics of the study participants are presented in Table 1. Men were slightly younger than women (mean age 69.1 vs 71.3 years, *p* = 0.009) and had higher waist circumference (102.3 vs 97.2 cm, *p* < 0.001). Men also reported heavy alcohol consumption more frequently than women (13.6% vs 6.2%, *p* = 0.014).

**Table 1.** Baseline characteristics of HeCES2 study cohort by sex.

|  | Men (n = 328) | Women (n = 155) |  |
| --- | --- | --- | --- |
| Age | 69.1 ± 8.2 | 71.1 ± 8.9 | p=0.016 |
| BMI | 27.4 ± 3.9 | 27.9 ± 5.7 | p=0.295 |
| Waist measurement | 102.4 ± 10.5 | 97.1 ± 14.3 | p<0.001 |
| Symptomatic plaque | 207 (63.1%) | 109 (70.3%) | p=0.120 |
| History of stroke or TIA | 151 (46.2%) | 56 (36.1%) | p=0.037 |
| Concomitant diseases |  |  |  |
| Kidney dysfunction | 53 (16.2%) | 24 (15.5%) | p=0.850 |
| Coronary artery disease (CAD) | 144 (43.9%) | 36 (23.2%) | p<0.001 |
| Hypertension | 268 (81.7%) | 125 (80.6%) | p=0.780 |
| Chronic heart failure | 32 (9.8%) | 19 (12.3%) | p=0.404 |
| Atrial fibrillation | 62 (18.9%) | 21 (13.5%) | p=0.145 |
| Peripheral artery disease | 62 (18.9%) | 23 (14.8%) | p=0.274 |
| Diabetes | 112 (34.1%) | 50 (32.3%) | p=0.682 |
| Smoking status | 124 (37.8%) | 59 (38.1%) | p=0.954 |
| Alcohol consumption | 44 (13.4%) | 9 (5.8%) | p=0.014 |
| Medications |  |  |  |
| Low-dose aspirin | 167 (50.9%) | 79 (51.0%) | p=0.991 |
| Clopidogrel | 47 (14.3%) | 28 (18.1%) | p=0.290 |
| Dual antiplatelet therapy (ASA+CLO) | 12 (3.7%) | 10 (6.5%) | p=0.169 |
| Lipid lowering agent | 317 (96.6%) | 148 (95.5%) | p=0.529 |
| ACEi or ATRb | 243 (74.1%) | 108 (69.7%) | p=0.310 |
| Beta blocker | 177 (54.1%) | 90 (58.1%) | p=0.417 |
| Calcium channel blocker | 113 (34.5%) | 61 (39.4%) | p=0.295 |
| Diabetes medication | 104 (31.7%) | 48 (31.0%) | p=0.870 |
| Lab measurements |  |  |  |
| Total cholesterol (mmol/L) | 5.7 ± 2.0 | 6.1 ± 1.2 | p=0.004 |
| LDL cholesterol (mmol/L) | 3.7 ± 1.0 | 3.9 ± 1.1 | p=0.015 |
| hs-CRP (mg/L) | 5.3 ± 13.1 | 5.5 ± 9.1 | p=0.866 |
| ApoB/ApoA | 0.6 ± 0.3 | 0.6 ± 0.3 | p=0.784 |
| P-Fibrinogen (g/L) | 3.9 ± 0.9 | 4.1 ± 0.9 | p=0.255 |

ASA+CLO, Low-dose aspirin and clopidogrel ACEi, Angiotensin-converting enzyme inhibitor ATRb, Angiotensin receptor blocker hs-CRP, high-sensitivity C-reactive protein ± 1 SD

Men more often had a history of stroke or transient ischemic attack (TIA) prior to carotid endarterectomy (46.2% vs 35.2%, *p* = 0.019) and had a higher prevalence of coronary artery disease (42.9% vs 24.1%, *p* < 0.001). Women had higher total and LDL cholesterol concentrations than men (total cholesterol 6.1 vs 5.6 mmol/L, *p* = 0.004; LDL cholesterol 3.9 vs 3.6 mmol/L, *p* = 0.015), whereas the ApoB/ApoA ratio did not differ between the sexes.

No statistically significant sex differences were observed in plaque symptom status or the distribution of neurological symptom types.

### Female carotid plaques exhibited greater calcification than male plaques

Female patients had a higher proportion of plaque calcification than male patients, reflected by higher median total calcification area (%) (median 15.8 % vs 10.8 %, p = 0.002), nodular calcification area (median 3.0 % vs 1.9 %, p = 0.030), and sheet calcification area (%) (median 9.3 % vs 5.3 %, p < 0.01) (Supplementary Figure S3). Histological total calcification area (%) correlated with the CTA-based calcification score (Spearman’s rho 0.56, 95% CI 0.50–0.62, p < 0.001; Supplementary Figure S1).

### Total and sheet calcification were inversely associated with plaque symptomatic status in the overall cohort and in men

In multivariable-adjusted logistic regression models, higher sheet calcification and total calcification were associated with lower odds of symptomatic presentation (Table S1). Per 10-% increase, the odds ratio was 0.79 (95% CI 0.67–0.92, p=0.004) for sheet calcification and 0.85 (95% CI 0.75–0.97, p=0.016) for total calcification. Nodular calcification was not associated with symptomatic status (OR 0.98, 95% CI 0.75–1.29, p=0.877). These inverse associations between plaque symptomatic status and sheet and total calcification were observed in men, whereas no corresponding associations were observed in women (Table S2). The sex-dependency of these associations was further confirmed by a sex interaction model for both sheet calcification (interaction OR 1.77, 95% CI 1.25–2.50, p=0.001) and total calcification (interaction OR 1.44, 95% CI 1.09–1.90, p=0.011) (Table S3).

### Stroke-associated plaques showed lower sheet calcification in men but not in women

In the overall cohort, plaques causing stroke had lower total calcification (%) than asymptomatic plaques (median 7.4 % vs 16.3 %, *p* < 0.001), an association driven by sheet calcification (median 4.1 % vs 9.3 %, *p* < 0.001) (Figures 2-3, Tables S5-S7). Plaque nodular calcification did not differ between these groups.

**Figure 1.**
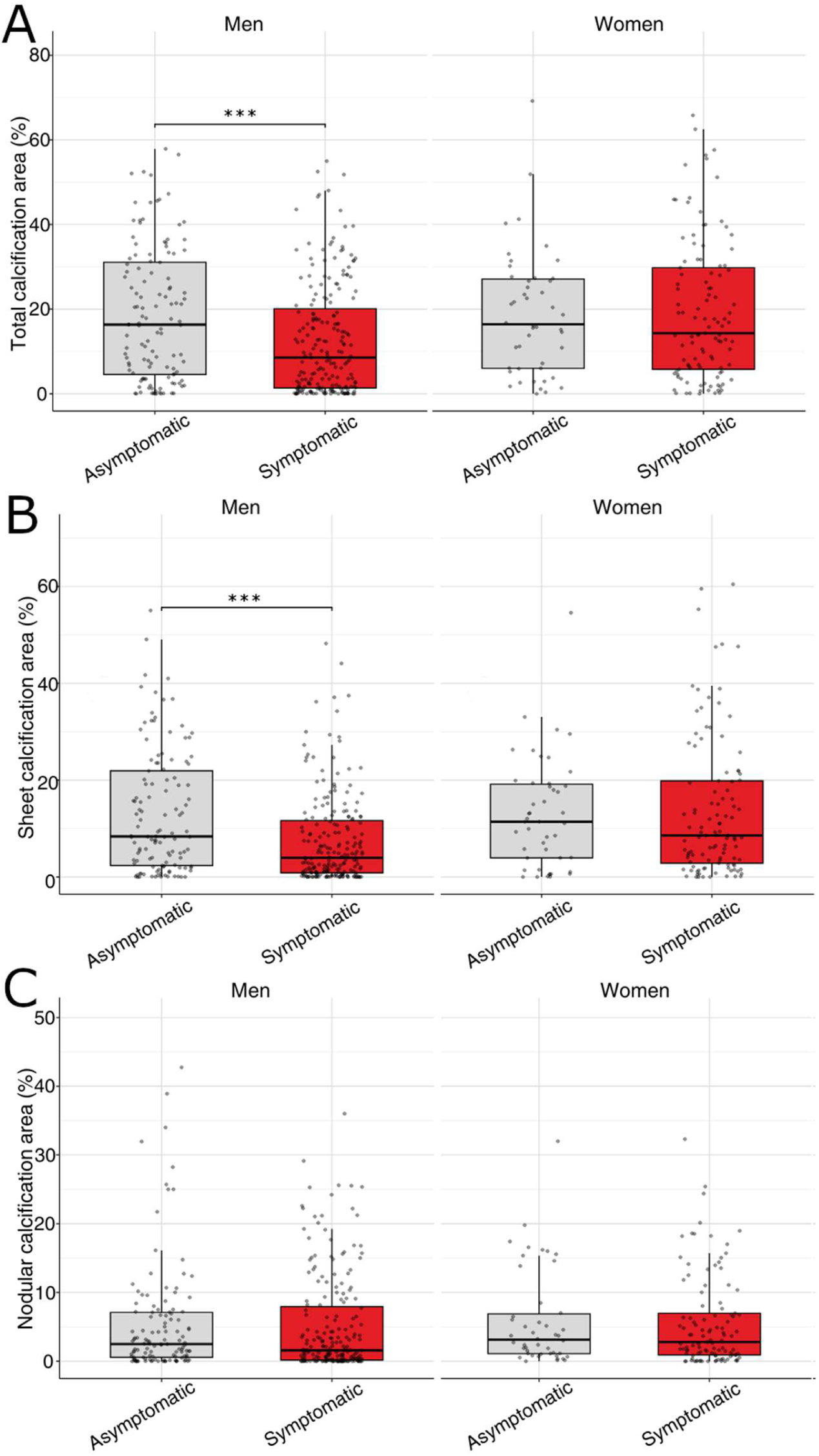
Carotid plaque (A) total calcification area, (B) sheet calcification area, and (C) nodular calcification area according to symptom status. Data are presented as box plots with dots representing individual measurements. Statistically significant differences between asymptomatic and symptomatic status are indicated by *p* < 0.05 (\**), p < 0.01 (**), and p < 0.001 (\*\*\**).

**Figure 2.**
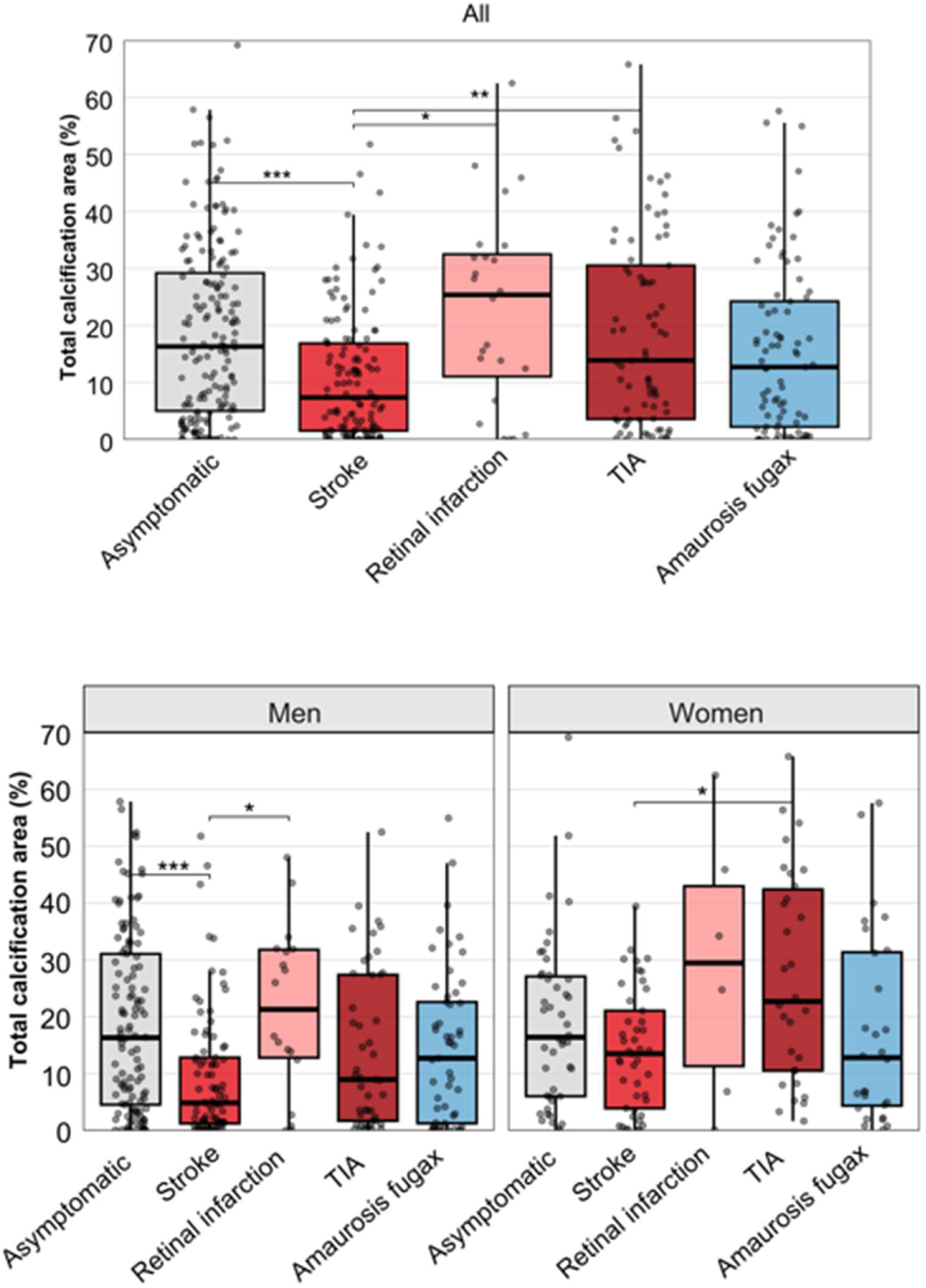
Plaque total calcification area (%) according to neurological symptom type. Data are presented as box plots with dots representing individual measurements. Statistically significant differences between neurological symptom types are indicated by *p* < 0.05 (\**), p < 0.01 (**), and p < 0.001 (\*\*\**).

**Figure 3.**
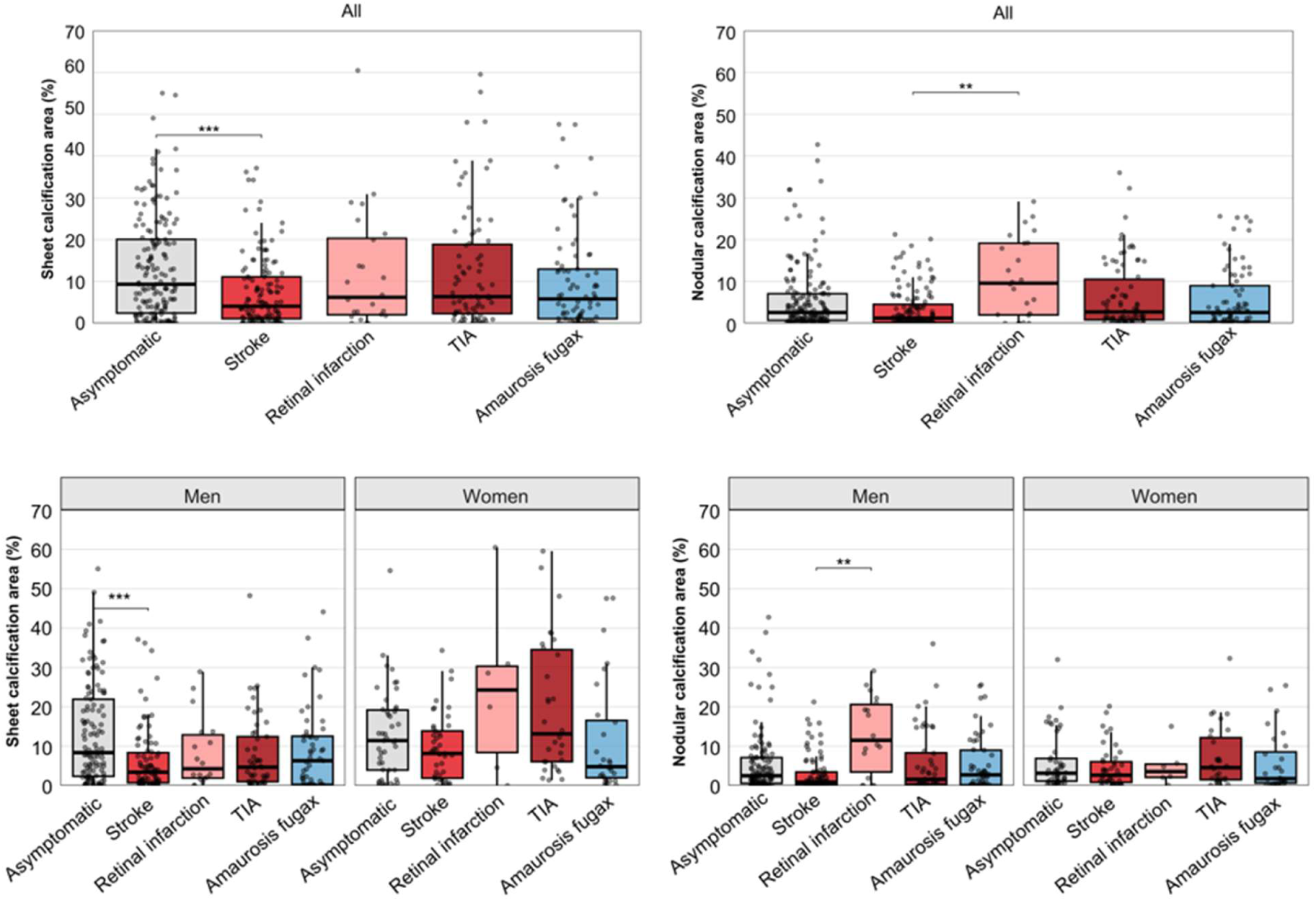
Plaque (A) sheet calcification area (%) and (B) nodular calcification area (%) according to neurological symptom type. Data are presented as box plots with dots representing individual measurements. Statistically significant differences between neurological symptom types are indicated by *p* < 0.05 (\**), p < 0.01 (**), and p < 0.001 (\*\*\**).

Sex-stratified analyses revealed that these differences were confined to men, in whom stoke-associated plaques were significantly less calcified than asymptomatic plaques (median total calcification 4.9 % vs 16.3 %, *p* < 0.001), predominantly due to reduced sheet calcification (median 3.4 % vs 8.4 %, *p* < 0.001). In women, no significant differences in calcification quantity or subtype were observed between asymptomatic and stroke plaques.

### High total calcification was associated with TIA in women

In the overall cohort, total calcification area (%) differed between plaques causing stroke and TIA (median 7.4 % vs 13.9 %, *p* = 0.003). Sex-stratified analyses demonstrated that this association was driven by women, in whom TIA plaques exhibited higher total calcification than stroke plaques (median 22.7 % vs 13.5 %, *p* = 0.024). Stroke and TIA plaques in women were similar in calcification subtype proportions.

### Retinal infarction was associated with higher total calcification and nodular calcification than stroke

Patients presenting with retinal infarction had higher total and nodular plaque calcification (%) than patients presenting with stroke (median total calcification 25.4 % vs 7.4 %, *p* = 0.003; median nodular calcification 9.6 % vs 1.2 %, *p* = 0.004). In sex-stratified analysis, this association was observed in men, in whom retinal infarction plaques exhibited higher total calcification than stroke plaques (median 21.3 % vs 4.9 %, *p* = 0.024), driven specifically by nodular calcification (median 11.5 % vs 0.85 %, *p* = 0.003). Due to the small number of female patients presenting with retinal infarction (n = 6), sex-stratified analyses could not be performed in women.

### CTA-based calcium score differed between patients according to symptom status

In ordinal regression analysis, radiological CTA-based calcium score differed significantly between neurological symptom types (Figure 4). Compared with asymptomatic patients, stroke was associated with lower odds of a higher calcium score (OR 0.48, 95% CI 0.30–0.77, p = 0.002), whereas retinal infarction was associated with higher odds (OR 2.54, 95% CI 1.05–6.31, p = 0.039). These associations were observed predominantly in men, among whom stroke was associated with lower odds (OR 0.44, 95% CI 0.25–0.78, p = 0.005) and retinal infarction with higher odds (OR 3.56, 95% CI 1.24–11.11, p = 0.021) of a higher radiological calcium score. No significant associations were observed among women.

**Figure 4.**
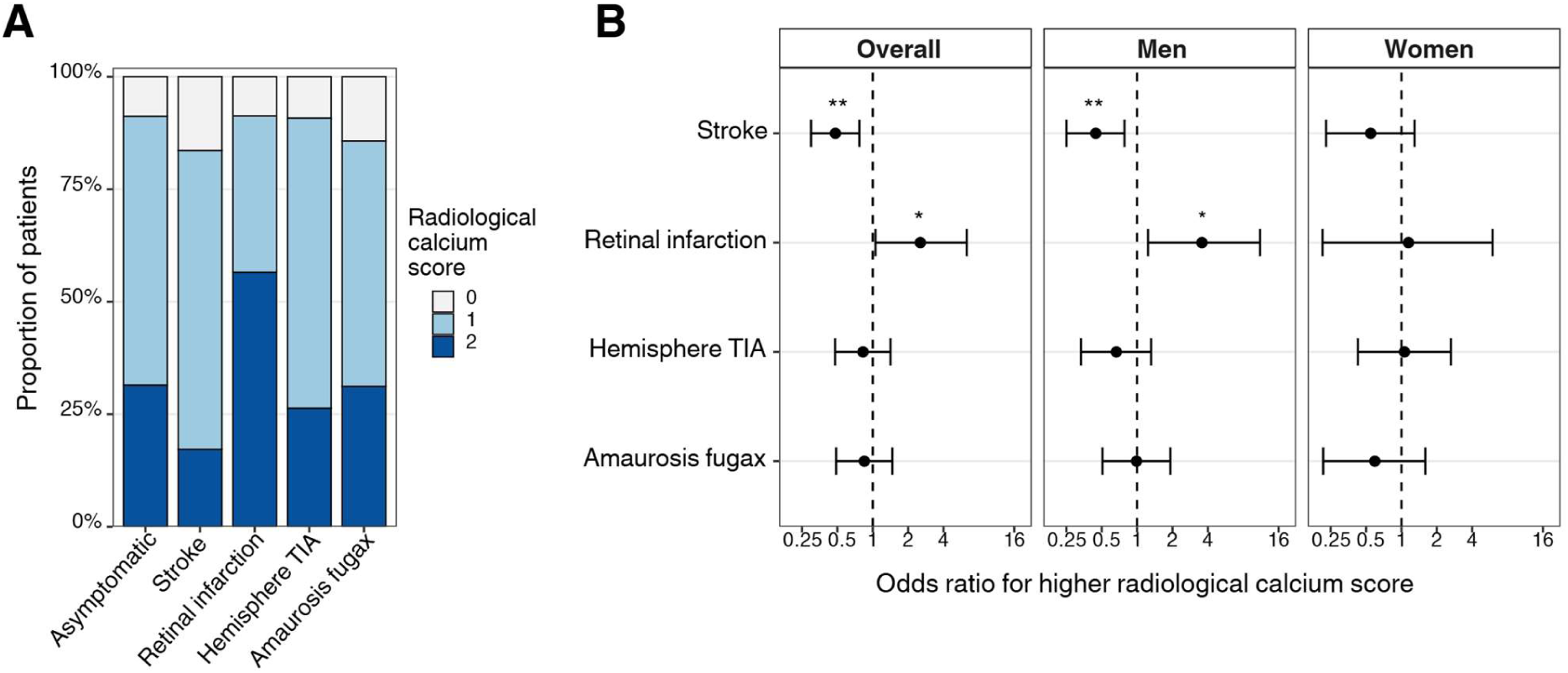
Distribution of radiological calcium score according to neurological symptom type and corresponding ordinal logistic regression results. (A) Proportional distribution of radiological calcium score categories by symptom type. (B) Odds ratios for belonging to a higher radiological calcium score category compared with asymptomatic patients, shown for the overall cohort and stratified by sex. Asymptomatic patients served as a reference group. Error bars indicate 95% CIs. Statistically significant associations are indicated by *p* < 0.05 (\**), p < 0.01 (**),* and *p < 0.001 (\*\*\**).

## Discussion

In this histological study of carotid plaques from patients undergoing carotid endarterectomy, plaque calcification showed distinct associations with cerebrovascular presentation according to calcification subtype and sex. In addition to the overall lower total and sheet calcification in symptomatic than asymptomatic plaques, three principal sex-dependent findings emerged. First, women had a higher relative plaque calcification burden than men. Second, in men, low sheet calcification characterized symptomatic and stroke-associated plaques, supporting sheet calcification as a marker of plaque stability. Third, nodular calcification was associated with retinal infarction rather than stroke. These findings suggest that carotid plaque calcification should not be interpreted as a uniform plaque feature, and that distinguishing between sheet and nodular calcification may provide clinically relevant insight into plaque biology and neurological presentation.

Previous studies on carotid plaque calcification have yielded partly conflicting results. Several imaging studies have suggested that calcification is associated with more stable plaque morphology and a lower likelihood of cerebrovascular ischemia,^7,8^ whereas other studies have linked spotty and rim sign calcification patterns to symptomatic disease.^9,28^ These discrepancies may partly reflect differences in study populations, imaging modalities, and definitions of calcification. Our findings extend this literature by showing that the clinical implications of calcification depend on its morphological subtype. In particular, sheet calcification was inversely associated with symptomatic plaque and stroke in men, whereas nodular calcification showed a distinct association with retinal infarction. Thus, total calcification alone may obscure biologically and clinically relevant differences between calcification phenotypes.

The inverse association between sheet calcification and stroke in men is consistent with the concept that large, confluent calcifications may represent a stabilizing plaque feature. Sheet calcification, by definition, consists of larger and more homogeneous calcium plates. In the present study, this interpretation was further supported by the inverse associations between calcification measures and vulnerable plaque features, including intraplaque haemorrhage and ulceration, particularly in men. These findings are in line with prior observations that plaques causing stroke tend to contain more unstable features, such as haemorrhage, lipid-rich necrotic core, and ulceration.^29–32^ Our results add to this literature by suggesting that low sheet calcification may be one histological feature of stroke-associated plaque vulnerability, especially in male patients.

The association between nodular calcification and retinal infarction provides a different perspective on plaque calcification. Previous histological work comparing ocular and cerebral ischemic events reported that plaques associated with ocular ischemia contained fewer vulnerable features and more calcification than plaques associated with cerebral events.^21^ In the present study, this association was further specified at the level of calcification morphology: retinal infarction was particularly associated with nodular calcification. Nodular calcification has been described as an irregular and potentially thrombus-associated form of calcification,^6,10^ but its clinical significance in carotid plaques has remained poorly understood. Our findings suggest that nodular calcification may not simply represent plaque stability or instability in the same manner as sheet calcification. Instead, it may reflect a distinct plaque phenotype associated with retinal ischemic symptoms. One possible explanation is that shed calcified microemboli preferentially enter the retinal circulation because of their small size or local flow conditions, or that they are more likely to cause clinically apparent symptoms in the retina, which has smaller vessels and more limited collateral circulation than the cerebral hemispheres.^33^ The limited number of women with retinal infarction prevented robust sex-stratified analysis of this event subtype, and this finding should therefore be validated in larger cohorts.

The present findings further support the view that cerebrovascular event subtypes are not biologically homogeneous. Stroke, TIA, retinal infarction, and amaurosis fugax are often grouped together as symptomatic carotid disease, but previous plaque studies have suggested that their underlying plaque features may differ.^21,34–35^ In our cohort, stroke-associated plaques in men were relatively less calcified, particularly with respect to sheet calcification, whereas plaques associated with TIA in women showed higher total calcification than stroke-associated plaques. Furthermore, women had a greater relative amount of plaque calcification than men overall, yet calcification was not associated with symptomatic plaque or stroke in women in the same way as in men. These findings are consistent with prior studies showing that carotid plaques from men more often exhibit vulnerable features^17,36^ and suggest that calcification may have different biological or clinical implications in women and men.

Several mechnisms may contribute to these observations. Interestingly, a study of microembolization during carotid artery stenting found that asymptomatic patients tended to produce a greater number of smaller, calcified particles, whereas larger, lipid-rich particles were more commonly observed in symptomatic patients.^37^ This may indicate that heavily calcified plaques are less prone to rupture or generate large emboli causing ischemic stroke, although this interpretation remains speculative. Alternatively, sex-specific differences in plaque composition, thromboinflammatory pathways, embolic potential, or downstream vascular susceptibility may influence whether carotid plaque disease manifests as stroke, TIA, or retinal ischemia. A contemporary estrogen-mediated explanation is unlikely, as the women in this cohort were predominantly postmenopausal. However, postmenopausal women may be more prone to developing local vessel wall thrombosis over small surface erosions because of increased platelet reactivity and procoagulant activity, including elevated levels of plasminogen activator inhibitor-1 (PAI-1) coagulation factors, particularly PAI-1.^38^ This could manifest as TIA without plaque rupture and large thromboembolism. This could result in smaller embolic events presenting as TIAs without overt plaque rupture or large thromboembolism. Other sex-related mechanisms, including differences in immune responses,^36^ vascular remodeling,^39^ coagulation,^40^ and calcification pathways^41^ may contribute to these divergent plaque phenotypes. Collectively, these findings emphasize the importance of considering both sex and symptom subtype when evaluating carotid plaque biology and underscore the need for sex-stratified analyses in studies of carotid atherosclerosis and plaque imaging.

Because histological plaque material is not routinely available in clinical practice, an important translational question is whether symptom-related differences in calcification can also be detected by routine imaging. We found that CTA-based radiological calcium score correlated with histological total calcification. Moreover, the imaging findings paralleled the histological results: lower radiological calcium score was associated with stroke, whereas higher calcium score was associated with retinal infarction, particularly in men. These findings support the potential clinical relevance of plaque calcification assessment by routine CTA. However, conventional CTA scoring primarily captures calcification burden and does not reliably distinguish sheet from nodular calcification. Emerging photon-counting CT and micro-CT approaches may eventually allow more detailed characterization of calcification morphology.^42–43^

Overall, our findings suggest that carotid plaque calcification may help explain why plaques with similar degrees of stenosis can lead to different clinical manifestations. Current decision-making for carotid endarterectomy is largely based on stenosis severity and symptom status; plaque composition is known to be a significant but less well understood feature of the biological risk. The present data indicate that calcification morphology, rather than calcification burden alone, may be relevant for understanding plaque behavior. In particular, low sheet calcification may indicate stroke-associated plaque vulnerability in men, whereas nodular calcification may identify a plaque phenotype linked to retinal infarction. These observations require validation but may inform future studies integrating histology, CTA-based plaque characterization, and sex-specific risk assessment.

This study has several strengths, including a well-characterized prospectively collected carotid endarterectomy cohort, detailed neurological and ophthalmologic symptom classification, histological assessment of excised plaques, and AI-based quantification of calcification subtypes. The use of automated image analysis enabled reproducible quantification of sheet and nodular calcification in a large histological dataset. Several limitations should also be acknowledged. This was a single-center, cross-sectional analysis of patients selected for carotid endarterectomy, and the findings may not be generalizable to patients with mild or medically managed carotid atherosclerosis. The number of patients with retinal infarction was limited, particularly among women, restricting sex-stratified analyses for this event subtype. Because multiple comparisons across symptom subtypes were performed, these findings should be interpreted as exploratory and require external validation. Finally, histological calcification was quantified from representative plaque sections and may not fully capture the three-dimensional calcification burden or spatial distribution of the entire plaque.

In conclusion, histologically defined carotid plaque calcification subtypes are associated with cerebrovascular presentation in a sex-dependent manner. Low sheet calcification characterized stroke-associated plaques in men, whereas nodular calcification was associated with retinal infarction. These findings support the biological and clinical relevance of distinguishing calcification morphology in carotid atherosclerosis and suggest that calcification subtype may help explain differences in neurological manifestations and risk-stratification among patients with carotid stenosis.

## Data Availability

The individual-level data underlying this article cannot be shared publicly or upon request because they contain sensitive patient information and disclosure is restricted by applicable data protection regulations and institutional data-governance requirements. Aggregated data supporting the findings are included in the article and its Supplemental Material.

## Acknowledgments

The authors thank all patients participating in the HeCES2 study. Authors also thank Heli Silvennoinen and Leena Valanne for for their valuable input in radiology.

## Sources of Funding

This study was supported by grants from Academy of Finland, Paavo Nurmi Foundation, the Paulo Foundation, Sigrid Jusélius Foundation, Päivikki and Sakari Sohlberg Foundation, Jane and Aatos Erkko foundation, The Finnish Foundation for Cardiovascular Research, Maud Kuistila memorial foundation, Juhani Aho Foundation for Medical Research, Helsinki University Hospital District governmental research funds and the Finnish Medical Foundation. Wihuri Research Institute is maintained by the Jenny and Antti Wihuri Foundation.

## Disclosures

JS reports lecturer honoraria from Abbott, Amgen, Lilly, Novo Nordisk, Novartis, and a grant from Finnish Cardiac foundation, Finnish Cultural foundation and Special Government grants, and consulting honoraria from Amarin, Amgen, GSK, Lilly, Novo Nordisk and Novartis Ltd (all unrelated to the present study).

No potential conflict of interest was reported by the other authors.

## Non-Standard Abbreviations and Acronyms

ACEi: angiotensin-converting enzyme inhibitor
AI: artificial intelligence
ASA: low-dose aspirin / acetylsalicylic acid
ATRb: angiotensin receptor blocker
BMI: body mass index
CAD: coronary artery disease
CEA: carotid endarterectom
CI: confidence interval
CLO: clopidogrel
CT: computed tomography
CTA: computed tomography angiography
ESVS: European Society of Vascular Surgery
HE: hematoxylin and eosin
HeCES2: Helsinki Carotid Endarterectomy Study 2
hs-CRP: high-sensitivity C-reactive protein
ICA: internal carotid artery
IPH: intraplaque hemorrhage
IQR: interquartile range
LDL: low-density lipoprotein
NASCET: North American Symptomatic Carotid Endarterectomy Trial
OR: odds ratio
SD: standard deviation
TIA: transient ischemic attack

## Notes

### Competing Interest Statement

The authors have declared no competing interest.

### Clinical Protocols

https://doi.org/10.1080/07853890.2018.1494851

### Author Declarations

The study was approved by the Ethics Committee of medicine of the Hospital District of Helsinki and Uusimaa.

